# Trends and Key Contributors of Racial and Ethnic Disparities in Life’s Essential 8: NHANES 2011-2020

**DOI:** 10.1101/2024.11.19.24317567

**Authors:** Huanhuan Yang, Chenxi Huang, Mitsuaki Sawano, Jeph Herrin, Kamil F. Faridi, Zhihui Li, Erica Spatz, Harlan M. Krumholz, Yuan Lu

## Abstract

**IMPORTANCE:** Significant racial and ethnic differences exist in Life’s Essential 8 (LE8), but the trends in these disparities over time are not well understood. Additionally, the key components of LE8 to these differences are unclear.

**OBJECTIVE:** To evaluate trends in racial and ethnic disparities in LE8 over a 10-year period and to identify the primary contributors among the eight LE8 components to these disparities.

**DESIGN, SETTING, AND PARTICIPANTS:** Serial population based cross-sectional study of the National Health and Nutrition Examination Survey (NHANES) from 2011 to 2020, that included 16,104 adults aged 20-79 years.

**EXPOSURE:** Self-reported race and ethnicity.

**MAIN OUTCOME AND MEASURES:** Trends in racial and ethnic differences in LE8, and primary contributors to the disparity.

**RESULTS:** The mean age of the participants was 46.2 (SE: 0.33) years; 51.1% were women; 68.2%, 10.9%, 15.7%, and 5.2% were White, Black, Latino/Hispanic, and Asian adults. From 2011-2020, Asian adults had the highest LE8 score (71.2), followed by White (67.7) and Latino/Hispanic (65.9) adults, and Black adults (62.0) had the lowest LE8 score. These racial/ethnic differences in LE8 overall score did not significantly change from 2011 to 2020 (all p value > 0.05). However, the differences in several individual components of LE8 changed significantly. For example, the Latino/Hispanic-White difference in sleep health score significantly enlarged, from −1.25 to −4.38, with a difference-in-difference of −3.12 (95% CI: −5.83, −0.42; p = 0.020). In 2017-2020, all but blood lipids and nicotine exposure were negative contributors (Z-scores < 0) for the Black-White difference; nicotine exposure was the key positive (Z-score =1.01), while physical activity was the key negative (Z-score = −1.01) contributor for the Latino/Hispanic-White difference; nicotine exposure (Z-score = 2.59) and diet (Z-score = 2.12) were the primary positive contributors for Asian-White difference.

**CONCLUSIONS AND RELEVANCE:** Racial and ethnic disparities in overall LE8 scores compared to White adults remained largely unchanged from 2011 to 2020. These disparities were driven by varying components across different racial and ethnic groups, emphasizing the need for targeted, group-specific interventions.

**Key Points:** *Question:* What are the trends in racial and ethnic disparities in Life’s Essential 8 (LE8) metrics and what are the primary contributors to it over the past decade?

*Findings:* Racial and ethnic disparities in overall LE8 remained stable, though some component gaps widened or narrowed. Black-White disparities were driven by factors beyond blood lipids and nicotine exposure, with physical activity as the primary negative contributor to Latino/Hispanic-White disparities, and diet and nicotine exposure as key positive contributors to Asian-White disparities.

*Meaning:* These findings highlight the need for tailored interventions, providing insights to meet each racial and ethnic group’s unique needs.

## Introduction

Significant racial and ethnic disparities persist in cardiovascular health (CVH), contributing to unequal burdens of cardiovascular diseases (CVD) across populations.^1,2^ CVD remains the leading cause of death in the United States, with disproportionate impacts on racial and ethnic minorities, such as Black, Hispanic, and Asian populations.^3–5^ Despite numerous public health efforts aimed at improving CVH, these disparities continue to drive inequities in health outcomes. As the American Heart Association (AHA) shifted from “Life’s Simple 7 (LS7)” to the more comprehensive “Life’s Essential 8 (LE8)” framework in 2022, a broader approach to health promotion and disease prevention has emerged, highlighting the need to examine how well we are addressing these disparities.^6,7^

Despite this shift in focus, there is a significant gap in our understanding of how racial and ethnic disparities in LE8 scores have evolved over the past decade. Previous studies have noted persistent disparities in CVH among different racial and ethnic groups, but they have not comprehensively assessed national trends of racial disparity in LE8 scores over time.^8–10^ Additionally, the relative contributions of each component of LE8—such as diet, physical activity, nicotine exposure, sleep, and metabolic factors—remain unclear. Without understanding which factors drive these disparities, it is difficult to develop targeted interventions that can reduce the gaps and improve health equity.

Understanding the trends of racial disparity in LE8 scores is critically important, as addressing health disparities is a national priority. Various public health measures, such as guidelines, policy interventions, and community outreach programs, have been implemented to reduce these inequities.^11,12^ However, it remains unclear whether these efforts have had a meaningful impact on closing racial and ethnic gaps in CVH. Given the continued high burden of CVD in minority populations and the recent introduction of the LE8 framework, this is an opportune moment to investigate how these disparities have changed and whether specific components of LE8 have improved or worsened over time.

To address this gap, our study utilized data from the National Health and Nutrition Examination Survey (NHANES) from 2011 to 2020 to assess trends in racial and ethnic disparities in Life’s Essential 8 (LE8) metrics and the relative contributions of each LE8 component to these disparities among U.S. adults.

## Methods

### Study Design

The NHANES is a national representative dataset provided by the US Centers for Disease Control and Prevention.^13^ This survey employs a complex multistage probability sampling design to collect health and nutrition data from a nationally representative sample of the civilian, non-institutionalized U.S. population. Detailed information can be found on its website: https://www.cdc.gov/nchs/nhanes/index.htm.

For the present study, we utilized continuous NHANES data spanning ten years of four cycles, covering 2011-2012 to 2017-2020. Each cycle collects data through home interviews and subsequent examinations at a mobile examination center (MEC). Except for variety of anthropometric measurements, biological samples including blood were also collected for laboratory tests. The NCHS Research Ethics Review Board approved all survey protocols for NHANES, and participants provided written informed consent before participating in NHANES.

### Study Participants

From 2011 to 2020, the continuous NHANES survey included a total of 45,462 participants. Of these, we included 23,570 adults aged 20 to 79 years and self-identified as non-Hispanic White, non-Hispanic Black, Latino/Hispanic, or Asian. Then we excluded those pregnant or lactating females at the time of examination (n = 425); and those having self-reported history of coronary heart disease (CHD), angina, heart attack, or stroke (n = 1,977). Individuals with missing values for a specific component were excluded (n = 5,064). Ultimately, 16,104 adults with complete data for all 8 components were included in the main analysis (**eFig.1**).

**Figure 1.**
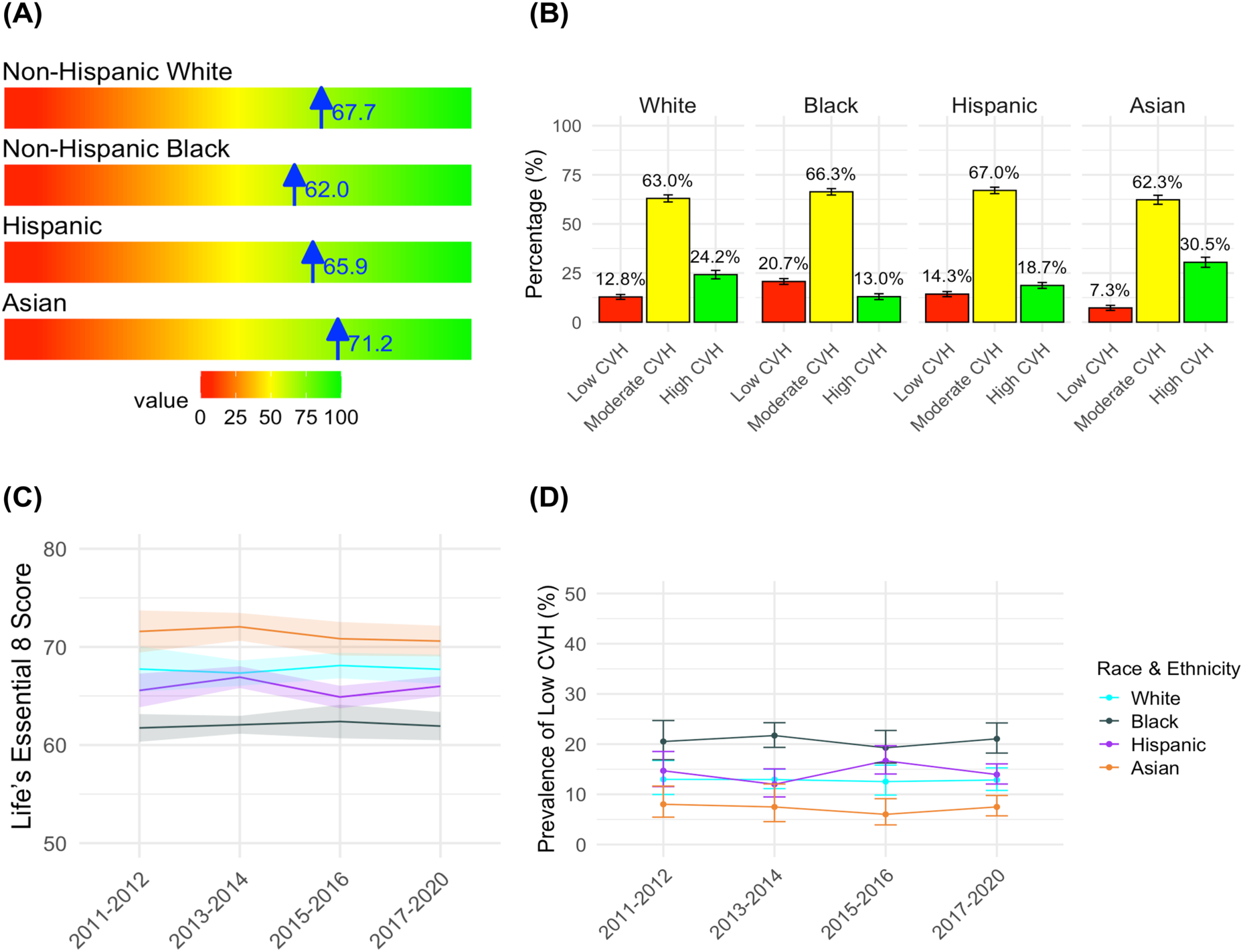
Distribution and Trends of Life’s Essential 8 Score and Cardiovascular Health Status. Figure (A) shows the survey weighted mean life’s essential 8 (LE8) scores across race and ethnicity groups; Figure (B) presents the survey weighted percentage of low cardiovascular health (CVH, LE8 score < 50), moderate CVH (LE8 score 50-79), and high CVH (LE8 score ≥ 80) with 95% confidence interval; Figure (C) depicts the trends of life’s essential 8 (LE8) scores by race and ethnicity; Figure (D) provides the weighted percentage with 95% confidence interval of participants with low cardiovascular health (LE8 score < 50) by race and ethnicity.

### LE8 Calculation

The LE8 score, derived from the AHA’s methods for quantifying CVH, encompasses eight components, each rated on a scale from 0 to 100.^7^ The scoring approach for quantifying LE8 were detailed in **eTable 1**. The assessment begins with a Dietary Approaches to Stop Hypertension (DASH)-style diet evaluated through the first 24-hour dietary recall, capturing intake from eight food and nutrient groups: fruits, whole grains, vegetables, nuts and legumes, low-fat dairy products, red and processed meats, sugar-sweetened beverages, and sodium. Each component is assigned between 1 to 5 points, accumulating up to a total of 40 points. Details on the DASH-style diet scoring are provided in elsewhere.^14^

Health behaviors, gathered from interviews, include self-reported weekly minutes of moderate to vigorous physical activity, nicotine exposure (including use of cigarettes and secondhand smoke exposure), and average hours of sleep per night. Anthropometric measurements considered are body mass index (BMI), calculated as body weight divided by height squared, and blood pressure, which is averaged across second to fourth measurements depending on availability.

Laboratory indicators assessed include non-high density lipoprotein cholesterol (non-HDL-C, equal to total cholesterol minus HDL-C), fasting blood glucose and casual Hemoglobin A1c (HbA1c) levels. The overall LE8 score is the mean of these eight metrics, also ranging from 0 to 100. Based on AHA recommendations, LE8 scores are categorized into three levels of cardiovascular health: low CVH (score < 50), medium CVH (score 50-79), and high CVH (score ≥ 80).^7^

### Covariates

Demographic and socioeconomic information was collected through interviews, including race, age, sex, the ratio of family income to poverty, education level, marital status, and insurance status (**eTable 1**). Race and ethnicity were self-reported and categorized as non-Hispanic White, non-Hispanic Black, Latino/Hispanic, and Asian. The ratio of family income to poverty was defined by the Department of Health and Human Services guidelines,^15^ calculated as the ratio of monthly family income to poverty levels and classified into four categories: low income (≤1.30), lower middle income (1.31-1.85), middle income (1.86-3.50), and high income (>3.50). Education levels were segmented into less than high school, high school graduate, and more than high school. Marital status was categorized as unmarried, and married or living with a partner. Insurance status was differentiated between uninsured and insured.

### Statistical Analysis

To incorporate the complex multistage sampling design of NHANES in the statistical analysis, sample weights for laboratory and physical examination (MEC weight) data were used to analyze the combined LE8 score. The data spanned from 2011 to 2020, with the period from 2017 to March 2020 considered as 3.2 pre-COVID years. For the analysis of each component, appropriate weights were chosen for each variable. Specifically, the dietary day one sample weight was used for the DASH diet; the interview weight was used for physical activity, nicotine exposure, and sleep health; and the MEC weight was used for blood pressure, BMI, blood glucose, and lipids. Age standardization was performed using the direct standardization method with 5-year intervals, based on the 2020 US Census population.^16^

To quantify racial and ethnic differences, we calculated the LE8 differences by subtracting the LE8 values of White adults from those of Black, Latino/Hispanic, and Asian adults for both the 2011-2012 and 2017-2020 survey cycles. We tested for an absolute difference in the racial and ethnic difference using a Z test. To assess the trends of racial and ethnic disparities over time, we employed a Difference-in-Differences (DiD) approach using survey-weighted regression models. A DiD analysis was conducted by including an interaction term between survey year and race and ethnicity group in the model, and the coefficients were extracted to evaluate the impact of time and race/ethnicity on the LE8 scores.

To evaluate the relative contributions of each component to the racial and ethnic disparities in LE8 scores, we calculated Z-scores for the differences with White adults across all components to account for differences in scale. Z-scores with positive or negative values were considered positive or negative contributors, respectively. The primary contributors were defined as the one or two components with the largest absolute Z-scores. A radar chart was then utilized to visualize and compare these relative contributions by race and ethnicity.

All analyses were performed using R software version 4.3.3, and all figures were plotted using the ggplot2 package. The “dietaryindex” package was used to calculate the DASH diet score.^17^ A two-sided p-value of less than 0.05 indicated a significant difference.

## Results

### Characteristics of Study Population

The analysis included 16,104 non-pregnant adults aged 20-79 years from the NHANES 2011-2020 dataset (**eFig.1**). The mean age of the study population was 46.2 years, with 51.1% of participants being women. In terms of racial and ethnic composition, 15.7% were Latino/Hispanic, 68.2% were White, 10.9% were Black, and 5.2% were Asian adults. Compared to White individuals, Latino/Hispanic, Black, and Asian individuals were generally younger. Additionally, Latino/Hispanic and Black individuals had lower income and education levels and were less likely to have health insurance (**Table 1**).

**Table 1.** Basic Characteristics by Race and Ethnicity in National Health and Nutrition Examination Survey (NHANES) 2011 to 2020 Years.

| Characteristics | Race and Ethnicity |  |  |  | *P* value |
| --- | --- | --- | --- | --- | --- |
|  | Non-Hispanic White | Non-Hispanic Black | Latino/Hispanic | Asian |  |
| Sample size | 6,068 | 3,918 | 4,144 | 1,974 |  |
| Population size | 145,325,656 | 23,162,248 | 33,533,230 | 11,067,640 |  |
| Age | 48.0±0.39 | 44.2±0.39 | 41.1±0.34 | 43.3±0.53 | <0.001 |
| Sex (n,%) |  |  |  |  | 0.001 |
| Male | 2,990 (48.91) | 1,876 (45.64) | 1,993 (51.37) | 983 (48.23) |  |
| Female | 3,078 (51.09) | 2,042 (54.36) | 2,151 (48.63) | 991 (51.77) |  |
| Ratio of family income to poverty (n,%) |  |  |  |  | <0.001 |
| ≤1.30 | 1,466 (13.88) | 1,260 (34.68) | 1,479 (38.09) | 310 (15.97) |  |
| 1.31–1.85 | 669 (8.14) | 482 (13.45) | 548 (14.24) | 181 (9.37) |  |
| 1.86–3.50 | 1,306 (23.83) | 905 (25.89) | 934 (25.49) | 418 (22.06) |  |
| >3.50 | 2,356 (54.15) | 917 (25.98) | 776 (22.18) | 935 (52.61) |  |
| Education (n,%) |  |  |  |  | <0.001 |
| Less than high school | 646 (7.14) | 606 (14.07) | 1,621 (33.40) | 212 (9.68) |  |
| High school | 1,383 (22.09) | 1,077 (27.79) | 899 (24.58) | 249 (12.59) |  |
| Greater than high school | 4,039 (70.77) | 2,234 (58.14) | 1,623 (42.02) | 1,512 (77.73) |  |
| Marital Status (n,%) |  |  |  |  | <0.001 |
| Unmarried | 2,226 (32.87) | 2,201 (56.52) | 1,435 (34.73) | 575 (30.52) |  |
| Married or living with partner | 3,842 (67.13) | 1,716 (43.48) | 2,706 (65.27) | 1,398 (69.48) |  |
| Insurance (n,%) |  |  |  |  | <0.001 |
| Uninsured | 924 (10.96) | 803 (22.18) | 1,400 (36.72) | 282 (12.92) |  |
| Insured | 5,140 (89.04) | 3,111 (77.82) | 2,733 (63.28) | 1,689 (87.08) |  |

Continuous variables were presented as survey-weighted means ± standard error (SE) and categorical variables as number of samples and survey-weighted percentage (n, %).

### Trends in Racial and Ethnic Differences in LE8 Scores

Overall, the highest mean LE8 score was observed among Asian individuals (71.2; 95% CI, 70.3-72.0), followed by White individuals (67.7; 95% CI, 66.9-68.6), Latino/Hispanic individuals (65.9; 95% CI, 65.2-66.6), and Black individuals (62.0; 95% CI, 61.3-62.7). The prevalence of low CVH was lowest among Asian individuals at 7.3% (95% CI, 6.0-8.5%), while Black individuals had the highest prevalence at 20.7% (95% CI, 19.2-22.2%) (**Figure 1A-B and eTable 2**).

Between 2011 and 2020, the racial and ethnic differences in LE8 scores did not show significant change (all p-values > 0.05). Black individuals consistently had lower scores than White individuals in both the 2011-2012 (Mean: −6.01; 95% CI, −8.72 to −3.29; p < 0.001) and 2017-2020 cycles (Mean: −5.79; 95% CI, −7.88 to −3.71; p < 0.001), with no significant change in the difference (DiD = 0.21; 95% CI, −3.38 to 3.80; p = 0.910). In contrast, Asian individuals consistently had higher LE8 scores than White individuals in both 2011-2012 (Mean: 3.83; 95% CI, 0.67 to 6.99; p = 0.018) and 2017-2020 cycles (Mean: 2.87; 95% CI, 0.70 to 5.05; p = 0.009), the gap was not significantly changed over time (DiD = −0.95; 95% CI, −4.05 to 2.15; p = 0.540) (**Figure 1B-C**, **Table 2**). No significant differences in LE8 scores were observed between Latino/Hispanic and White individuals in either the 2011-2012 or 2017-2020 survey cycles.

**Table 2.**
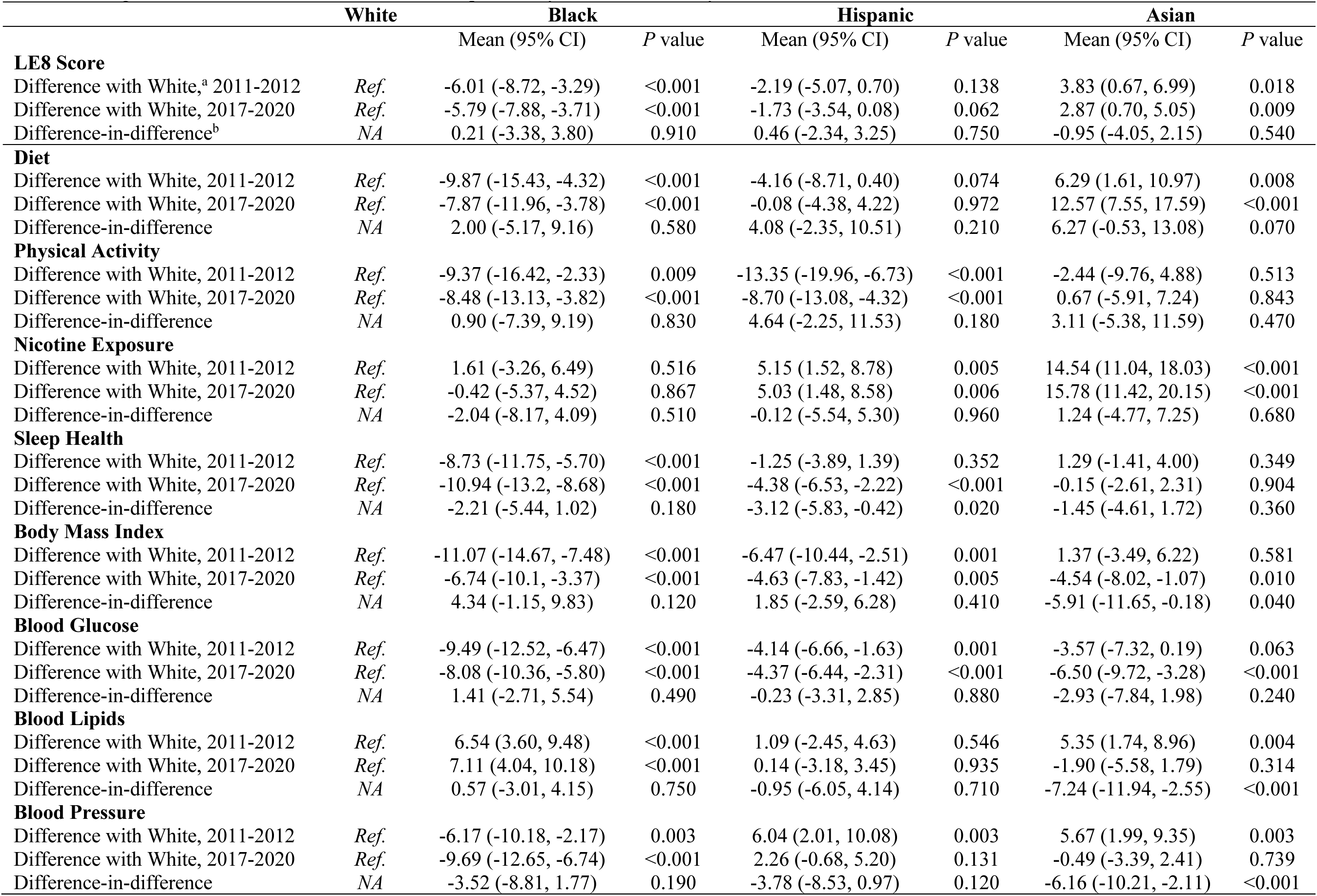
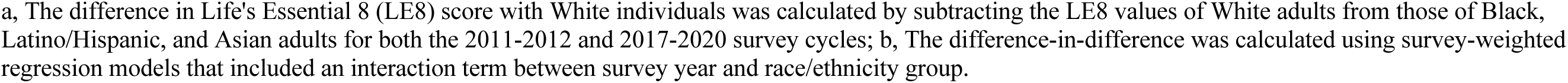
Changes in Life’s Essential 8 Score and Its Components by Race and Ethnicity.

|  | **White** | **Black** |  | **Hispanic** |  | **Asian** |  |
| --- | --- | --- | --- | --- | --- | --- | --- |
|  |  | Mean (95% CI) | P value | Mean (95% CI) | P value | Mean (95% CI) | P value |
| **LE8 Score** |  |  |  |  |  |  |  |
| Difference with White,<sup>a</sup> 2011-2012 | *Ref.* | -6.01 (-8.72, -3.29) | <0.001 | -2.19 (-5.07, 0.70) | 0.138 | 3.83 (0.67, 6.99) | 0.018 |
| Difference with White, 2017-2020 | *Ref.* | -5.79 (-7.88, -3.71) | <0.001 | -1.73 (-3.54, 0.08) | 0.062 | 2.87 (0.70, 5.05) | 0.009 |
| Difference-in-difference<sup>b</sup> | *NA* | 0.21 (-3.38, 3.80) | 0.910 | 0.46 (-2.34, 3.25) | 0.750 | -0.95 (-4.05, 2.15) | 0.540 |
| **Diet** |  |  |  |  |  |  |  |
| Difference with White, 2011-2012 | *Ref.* | -9.87 (-15.43, -4.32) | <0.001 | -4.16 (-8.71, 0.40) | 0.074 | 6.29 (1.61, 10.97) | 0.008 |
| Difference with White, 2017-2020 | *Ref.* | -7.87 (-11.96, -3.78) | <0.001 | -0.08 (-4.38, 4.22) | 0.972 | 12.57 (7.55, 17.59) | <0.001 |
| Difference-in-difference | *NA* | 2.00 (-5.17, 9.16) | 0.580 | 4.08 (-2.35, 10.51) | 0.210 | 6.27 (-0.53, 13.08) | 0.070 |
| **Physical Activity** |  |  |  |  |  |  |  |
| Difference with White, 2011-2012 | *Ref.* | -9.37 (-16.42, -2.33) | 0.009 | -13.35 (-19.96, -6.73) | <0.001 | -2.44 (-9.76, 4.88) | 0.513 |
| Difference with White, 2017-2020 | *Ref.* | -8.48 (-13.13, -3.82) | <0.001 | -8.70 (-13.08, -4.32) | <0.001 | 0.67 (-5.91, 7.24) | 0.843 |
| Difference-in-difference | *NA* | 0.90 (-7.39, 9.19) | 0.830 | 4.64 (-2.25, 11.53) | 0.180 | 3.11 (-5.38, 11.59) | 0.470 |
| **Nicotine Exposure** |  |  |  |  |  |  |  |
| Difference with White, 2011-2012 | *Ref.* | 1.61 (-3.26, 6.49) | 0.516 | 5.15 (1.52, 8.78) | 0.005 | 14.54 (11.04, 18.03) | <0.001 |
| Difference with White, 2017-2020 | *Ref.* | -0.42 (-5.37, 4.52) | 0.867 | 5.03 (1.48, 8.58) | 0.006 | 15.78 (11.42, 20.15) | <0.001 |
| Difference-in-difference | *NA* | -2.04 (-8.17, 4.09) | 0.510 | -0.12 (-5.54, 5.30) | 0.960 | 1.24 (-4.77, 7.25) | 0.680 |
| **Sleep Health** |  |  |  |  |  |  |  |
| Difference with White, 2011-2012 | *Ref.* | -8.73 (-11.75, -5.70) | <0.001 | -1.25 (-3.89, 1.39) | 0.352 | 1.29 (-1.41, 4.00) | 0.349 |
| Difference with White, 2017-2020 | *Ref.* | -10.94 (-13.2, -8.68) | <0.001 | -4.38 (-6.53, -2.22) | <0.001 | -0.15 (-2.61, 2.31) | 0.904 |
| Difference-in-difference | *NA* | -2.21 (-5.44, 1.02) | 0.180 | -3.12 (-5.83, -0.42) | 0.020 | -1.45 (-4.61, 1.72) | 0.360 |
| **Body Mass Index** |  |  |  |  |  |  |  |
| Difference with White, 2011-2012 | *Ref.* | -11.07 (-14.67, -7.48) | <0.001 | -6.47 (-10.44, -2.51) | 0.001 | 1.37 (-3.49, 6.22) | 0.581 |
| Difference with White, 2017-2020 | *Ref.* | -6.74 (-10.1, -3.37) | <0.001 | -4.63 (-7.83, -1.42) | 0.005 | -4.54 (-8.02, -1.07) | 0.010 |
| Difference-in-difference | *NA* | 4.34 (-1.15, 9.83) | 0.120 | 1.85 (-2.59, 6.28) | 0.410 | -5.91 (-11.65, -0.18) | 0.040 |
| **Blood Glucose** |  |  |  |  |  |  |  |
| Difference with White, 2011-2012 | *Ref.* | -9.49 (-12.52, -6.47) | <0.001 | -4.14 (-6.66, -1.63) | 0.001 | -3.57 (-7.32, 0.19) | 0.063 |
| Difference with White, 2017-2020 | *Ref.* | -8.08 (-10.36, -5.80) | <0.001 | -4.37 (-6.44, -2.31) | <0.001 | -6.50 (-9.72, -3.28) | <0.001 |
| Difference-in-difference | *NA* | 1.41 (-2.71, 5.54) | 0.490 | -0.23 (-3.31, 2.85) | 0.880 | -2.93 (-7.84, 1.98) | 0.240 |
| **Blood Lipids** |  |  |  |  |  |  |  |
| Difference with White, 2011-2012 | *Ref.* | 6.54 (3.60, 9.48) | <0.001 | 1.09 (-2.45, 4.63) | 0.546 | 5.35 (1.74, 8.96) | 0.004 |
| Difference with White, 2017-2020 | *Ref.* | 7.11 (4.04, 10.18) | <0.001 | 0.14 (-3.18, 3.45) | 0.935 | -1.90 (-5.58, 1.79) | 0.314 |
| Difference-in-difference | *NA* | 0.57 (-3.01, 4.15) | 0.750 | -0.95 (-6.05, 4.14) | 0.710 | -7.24 (-11.94, -2.55) | <0.001 |
| **Blood Pressure** |  |  |  |  |  |  |  |
| Difference with White, 2011-2012 | *Ref.* | -6.17 (-10.18, -2.17) | 0.003 | 6.04 (2.01, 10.08) | 0.003 | 5.67 (1.99, 9.35) | 0.003 |
| Difference with White, 2017-2020 | *Ref.* | -9.69 (-12.65, -6.74) | <0.001 | 2.26 (-0.68, 5.20) | 0.131 | -0.49 (-3.39, 2.41) | 0.739 |
| Difference-in-difference | *NA* | -3.52 (-8.81, 1.77) | 0.190 | -3.78 (-8.53, 0.97) | 0.120 | -6.16 (-10.21, -2.11) | <0.001 |

a, The difference in Life's Essential 8 (LE8) score with White individuals was calculated by subtracting the LE8 values of White adults from those of Black, Latino/Hispanic, and Asian adults for both the 2011-2012 and 2017-2020 survey cycles; b, The difference-in-difference was calculated using survey-weighted regression models that included an interaction term between survey year and race/ethnicity group.

### Trends in Racial and Ethnic Differences in Individual Components of LE8

Significant racial and ethnic differences were observed across several individual components of the LE8 score during the study period (**Figure 2, eTable 3**). Black individuals scored lower on most LE8 components compared to White individuals, except for blood lipids and nicotine exposure in 2011-2012. Notably, the Black-White gap in LE8 components did not significantly change from 2011 to 2020 for any component (**Table 2**, all p-values for DiD > 0.05).

**Figure 2.**
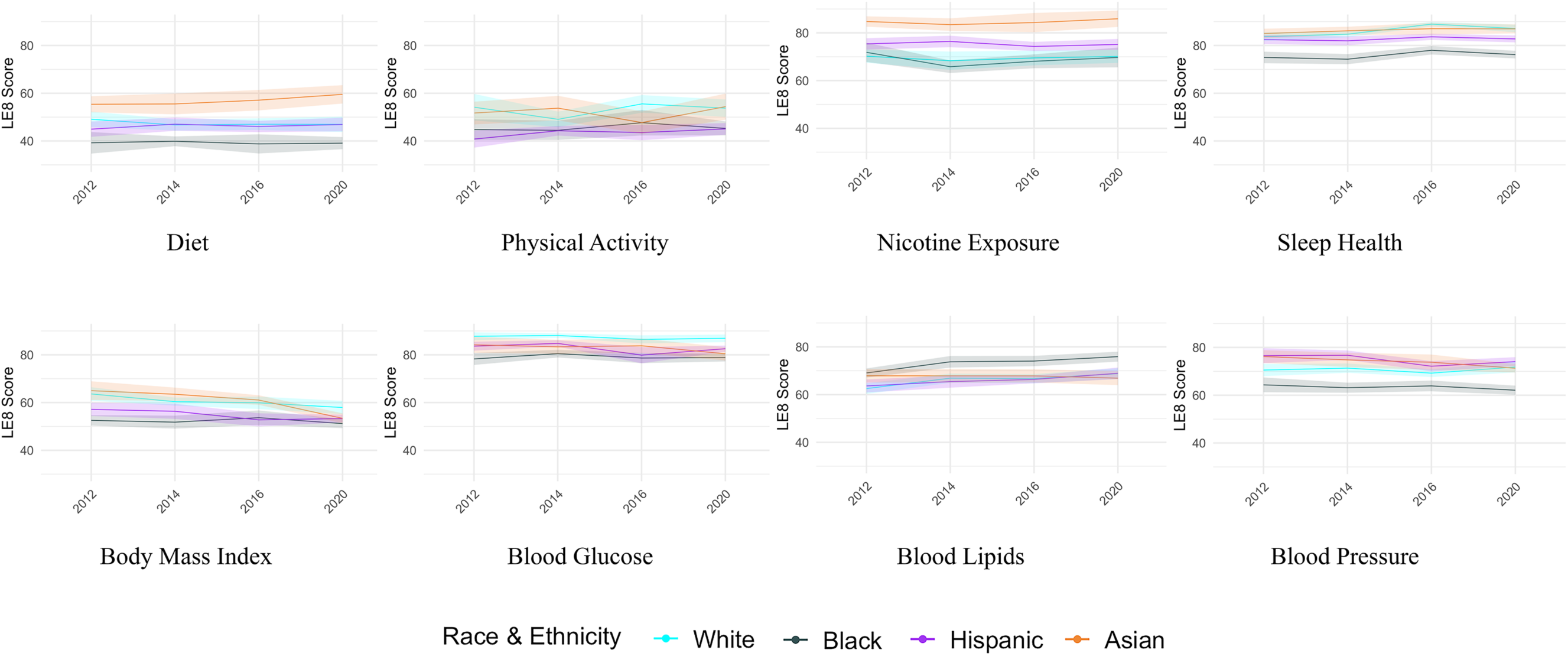
Trends of Each Component in Life’s Essential 8 Score by Race and Ethnicity.

Latino/Hispanic individuals had significantly lower physical activity, BMI, and blood glucose scores than White individuals in 2011-2012, but higher nicotine exposure and blood pressure scores. These disparities remained stable over time, except for sleep health, where the gap widened significantly from 2011-2012 to 2017-2020 (DiD = −3.12; 95% CI, −5.83 to −0.42; p = 0.020).

Asian individuals had higher scores in several LE8 components compared to White individuals in the 2011-2012 survey cycle, including diet, nicotine exposure, blood lipids, and blood pressure. Over time, the Asian-White differences in blood lipids (DiD = −7.24; 95% CI, −11.94 to −2.55; p < 0.001) and blood pressure (DiD = −6.16; 95% CI, −10.21 to −2.11; p < 0.001) narrowed significantly and were no longer significant by the 2017-2020 cycle. Furthermore, BMI score disparities reversed from being non-significant in 2011-2012 to significantly lower among Asian individuals compared to White individuals in 2017-2020 (DiD = −5.91; 95% CI, −11.65 to −0.18; p = 0.040) (**Table 2**).

### Key Contributors to Racial/Ethnic Differences in LE8 Scores

In the Black-White disparity, most LE8 components were negative contributors to the LE8 score gap in 2011-2012, with the exceptions of blood lipids and nicotine exposure. These relative contributions remained largely unchanged in 2017-2020 (**Figure 3, eTable 4**). For Latino/Hispanic-White disparities, key contributors in 2011-2012 included nicotine exposure and blood pressure (positive contributors) and physical activity (a negative contributor). By 2017-2020, the relative advantage in blood pressure had diminished, while other factors remained consistent. For Asian individuals, nicotine exposure was the strongest positive contributor to higher LE8 scores in 2011-2012, while physical activity and blood glucose were negative contributors. By 2017-2020, diet also became a major positive contributor alongside nicotine exposure; however, the advantages in BMI, blood lipids, and blood pressure had diminished or even reversed.

**Figure 3.**
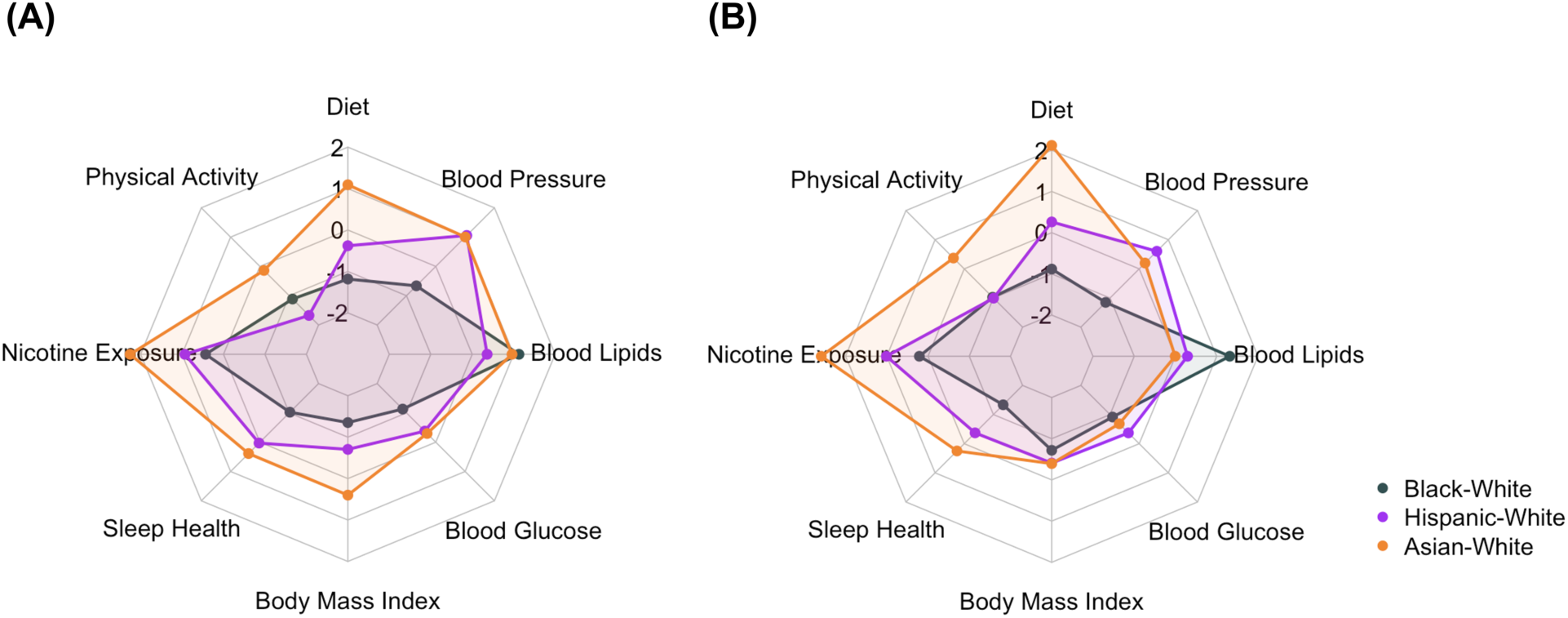
Relative Contributions of Each Component in Life’s Essential 8 to Racial and Ethnic Disparities. The difference in each component of Life’s Essential 8 (LE8) score compared to White individuals was calculated by subtracting the LE8 values of White adults from those of Black, Latino/Hispanic, and Asian adults for both the 2011-2012 (A) and 2017-2020 (B) survey cycles. Z-scores were obtained by standardizing the values of all components across all racial and ethnic groups.

## Discussion

This study examined trends in racial and ethnic differences in LE8 scores among American adults from 2011 to 2020, along with the key components contributing to these disparities. Our results revealed that Asian adults consistently had the highest LE8 scores and lowest rates of low CVH, while Black adults had the lowest LE8 scores and highest rates of low CVH. These disparities remained stable over the decade, with no significant overall improvement. However, we found evolving trends in individual LE8 components, including declines in BMI, blood lipids, and blood pressure scores among Asian individuals, and a widening gap in sleep health between Latino/Hispanic and White individuals. Black-White differences in LE8 scores remained stable, with the gap largely driven by components other than blood lipids and nicotine exposure.

Our study extends the prior literature in three important ways. First, while previous research has consistently shown significant racial and ethnic disparities in cardiovascular health,^18,19^ our study focuses on the updated LE8 framework, offering a more comprehensive analysis of CVH that includes newer components such as sleep health.^7^ Importantly, we analyzed trends in racial disparities in cardiovascular health using a statistical approach, demonstrating that these disparities persisted throughout the decade, particularly among Black adults, supplementing findings from prior studies.^8,9^

Second, unlike most previous studies that focused on overall CVH or limited components of health,^5,18,19^ we provide a detailed breakdown of racial and ethnic disparities across all LE8 components. Our findings expand on previous studies by thoroughly analyzing the racial and ethnic disparities for each LE8 component, revealing a worsening sleep health gap in the Latino/Hispanic population compared to White individuals. Additionally, the observed reversal in BMI advantages and the diminishing differences in blood lipids and blood pressure among Asian individuals provide a more detailed understanding of how health disparities manifest across various aspects of cardiovascular health.

Third, we are the first to assess the relative contribution of each LE8 component to overall racial and ethnic disparities, identifying specific areas where targeted interventions may be most effective. While socioeconomic factors have been citedas drivers of these disparities,^20,21^ our analysis goes further by quantifying the impact of each LE8 component on the observed differences, with particular emphasis on the roles of physical activity in Latino/Hispanic-White disparities, and diet and nicotine exposure in Asian-White disparities. This provides a more granular understanding compared to prior studies, which often treated these components collectively.

Our findings have significant implications for public health and clinical practice. Despite extensive public health efforts to address health disparities, including healthcare reforms like the Affordable Care Act,^22,23^ racial and ethnic gaps in CVH have not markedly improved over the past decade. This highlights the need for more effective, tailored interventions that target specific drivers of these disparities. For example, Black-White disparities are driven by multiple factors, including diet, physical activity, and sleep health, suggesting that culturally tailored outreach, enhanced access to preventive services, and behavioral support could be more effective than a one-size-fits-all approach. Similarly, the decline in health indicators like BMI, blood lipids, and blood pressure among Asian individuals calls for early, culturally appropriate interventions to prevent further deterioration. The widening sleep health gap among Latino/Hispanic individuals also underscores the need to prioritize underrecognized contributors to CVH, such as sleep, in public health initiatives. Addressing sleep health disparities may involve tackling broader social determinants like work conditions and stress,^24^ which disproportionately affect Latino/Hispanic communities. Public health strategies need to be adapted to meet the unique needs of each racial and ethnic group.

Our study has several limitations. First, behavioral variables such as diet, physical activity, nicotine exposure, and sleep health were self-reported, which may introduce bias. However, these measures have been widely used in previous research. Second, fasting plasma glucose measurements were only available for a subset of participants, limiting the generalizability of findings related to blood glucose. Finally, while NHANES employs strategies to reduce bias from nonresponse, residual effects may persist. To mitigate this, we adjusted for survey design and applied sample weights to account for nonresponse bias.

In conclusion, this study underscores the persistent racial and ethnic disparities in LE8 scores over the past decade. Targeted, effective interventions are necessary to address the key factors driving these differences and improve cardiovascular health equity.

## Data Availability

All data produced are available online at https://wwwn.cdc.gov/nchs/nhanes/Default.aspx

