## Supplemental tables and figures for "Trends and Key Contributors of Racial and Ethnic Disparities in Life’s Essential 8: NHANES 2011-2020"

**eFigure 1. Flow chart of participants.**

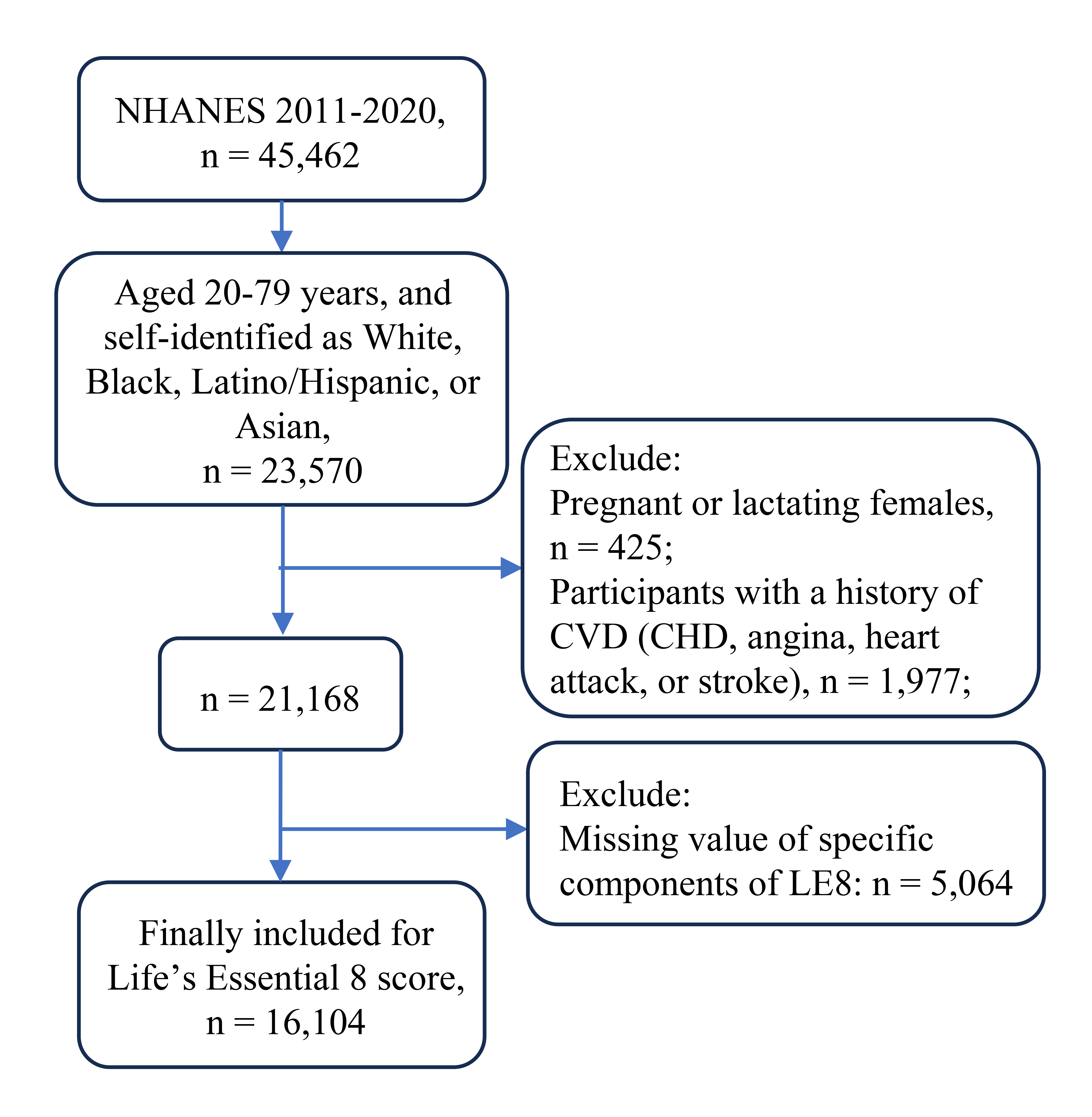

**eTable 1. Definition of Sociodemographic and Scoring Approach for Quantifying Life’s Essential 8 in the National Health and Nutrition Examination Surveys, 2011-2020.**

| **Risk factor** | **Ascertainment in NHANES** | **Definition of variables used in analysis** |
| --- | --- | --- |
| Ratio of family income to poverty | In-person interview: Please describe your family income (reported as a range value in dollars) | Ratio of family income to poverty was defined by the Department of Health and Human Services guidelines, was calculated as the ratio of monthly family income to poverty levels and classified into four categories: low income (≤1.30), lower middle income (1.31-1.85), middle income (1.86-3.50), and high income (>3.50). |
| Education level | In-person interview: What is the highest grade or level of school you have completed or the highest degree you have received? | Highest education level is classified as less than high school, high school, greater than high school. |
| Marital status | In-person interview: Please describe your current marital status. | Marital status is classified as married/ living with partners vs. unmarried (widowed, divorced, or separated; and never married). |
| Health insurance | In-person interview: Are you covered by health insurance or some other kind of health care plan? [Include health insurance obtained through employment or purchased directly as well as government programs like Medicare and Medicaid that provide medical care or help pay medical bills.] | Individuals are classified as insured if they had any private health insurance, Medicare, Medicaid, military plan, government or state-sponsored health plan. |
| DASH Diet | 24-h recall dietary interview (day 1 and day 2): The first dietary recall interview is collected in-person in the Mobile Examination Center (MEC) and the second interview is collected by telephone 3 to 10 days later. | DASH diet score was calculated using the method of Frank B. H. et al.;  **Scoring (Population):**  Points Quantile  100 ≥95^th^ %ile (top/ideal diet)  80 75^th^ – 94^th^ %ile  50 50^th^ – 74^th^ %ile  25 25^th^ – 49^th^ %ile  0 1^st^ – 24^th^ %ile (bottom/least ideal quartile) |
| Physical activity | In-person interview:  • Behavioral Risk Factor Surveillance Survey (BRFSS) physical activity instrument.  • Patients were asked if they participated in moderate or vigorous physical activity during the past 30 days. If they answered yes to either question, they were then asked the duration and frequency of their participation in physical activity for an average week. | Physical activity is calculated into recreational activities minutes  per week: (Days moderate recreational activities in a typical week * Minutes moderate recreational activities on a typical day) + (Days vigorous recreational activities in a typical week * Minutes vigorous recreational activities on a typical day).  **Metric:** Minutes of moderate (or greater) intensity activity per week  **Scoring:**  Points Minutes  100 ≥150  90 120 – 149  80 90 – 119  60 60 – 89  40 30 – 59  20 1 – 29  0 0 |
| Nicotine exposure | In-person interview:  • Ever smoke cigarettes in entire life  • Do you now smoke cigarettes?  • How long has it been since you quit smoking cigarettes?  • How many people who live here smoke cigarettes, cigars, little cigars, pipes, water pipes, hookah, or any other tobacco product? | Never smokers are defined as individuals who stated never smokes.  Former smokers are defined as individuals who stated that they had smoked but now did not smoke.  Current smokers are defined as individuals who stated that they smoked cigarettes currently.  Living with active indoor smoker in home are defined as live with at least one people who smoke any tobacco product.  **Metric:** Combustible tobacco use and/or inhaled nicotine delivery systems (NDS) use; or secondhand smoke exposure  **Scoring:**  Points Status  100 Never smoker  75 Former smoker, quit ≥ 5 yrs  50 Former smoker, quit 1 to < 5 yrs  25 Former smoker, quit <1 year, or currently  using inhaled NDS  0 Current smoker  Subtract 20 points (unless score is 0) for living with active indoor smoker in home. |
| Sleep health | In-person interview:  Number of hours usually sleep on weekdays or workdays. | **Metric:** Average hours of sleep per night  **Scoring:**  Points Level  100 7 to < 9 h  90 9 to < 10 h  70 6 to < 7 h  40 5 to < 6 or ≥ 10 h  20 4 to <5 h  0 <4 h |
| Body mass index (kg/m^2^) | Examination measurement:  Weight is measured on all examinees  Standing height is measured on all examinees 2 years and older | Body mass index (BMI, kg/m^2^) are calculated as body weight (kg) divided by height squared (m^2^).  **Metric:** BMI (kg/m^2^)  **Scoring:**  Points Level  100 <25  70 25.0 to 29.9  30 30.0 to 34.9  15 35.0 to 39.9  0 ≥ 40 |
| Blood glucose & Diabetes | In-person interview:  • Are you now taking diabetic pills to lower your blood sugar? These are sometimes called oral agents or oral hypoglycemic agents.  • Are you now taking insulin?  • Self-reported use of prescription medications during a one-month period prior to the survey date.  Lab measurement:  All participants ages 12 and older are given the option of a HbA1C% test during their physical examination. Besides, Participants aged 12 years and older who were examined in the morning session were tested for fasting glucose. | Diabetes is defined based on fasting glucose ≥ 126 mg/dL, or HbA1c ≥ 6.5%, or currently on antidiabetic medication.  **Metric:** Fasting blood glucose (FBG, mg/dL) or Hemoglobin A1c (%)  **Scoring:**  Points Level  100 No history of diabetes and FBG <100 (or  HbA1c < 5.7%)  60 No diabetes and FBG 100 – 125 mg/dL (or HbA1c 5.7-6.4%) (Pre-diabetes)  40 Diabetes with HbA1c <7.0%  30 Diabetes with HbA1c 7.0 – 7.9%  20 Diabetes with HbA1c 8.0 – 8.9%  10 Diabetes with Hb A1c 9.0 – 9.9%  0 Diabetes with HbA1c ≥10.0% |
| Blood lipids | Lab measurement:  All participants aged 6 years and older. | Non-HDL-cholesterol (mg/dL) was calculated by plasma total minus HDL-cholesterol;  **Metric:** Non-HDL-cholesterol (mg/dL)  **Scoring**:  Points Level  100 <130  60 130 – 159  40 160 – 189  20 190 – 219  0 ≥220  If drug-treated level, subtract 20 points |
| Blood pressure & Hypertension | In-person interview:  Are you now taking prescribed medicine for high blood pressure (BP)?  Examination measurement:  Blood pressure (BP) is measured on all examinees 8 years and older. | Hypertension is defined as average systolic blood pressure ≥ 140, or averaged diastolic blood pressure ≥ 90, or currently on antihypertensive medication.  **Metric:** Systolic and diastolic blood pressure (mm Hg)  **Scoring:**  Points Level  100 <120/<80 (Optimal)  75 120-129/<80 (Elevated)  50 130-139 or 80-89 (Stage I HTN)  25 140-159 or 90-99  0 ≥160 or ≥100  Subtract 20 points (unless score is 0) if treated level |

**eTable 2.** **Overall Life's Essential 8 Scores and Scores by Race and Ethnicity.**

|  | **Total** | **White** | **Black** | **Hispanic** | **Asian** | ***P*** **value** |
| --- | --- | --- | --- | --- | --- | --- |
| **Le8 score** |  |  |  |  |  |  |
| Total | 67.0 (66.4, 67.6) | 67.7 (66.9, 68.6) | 62.0 (61.3, 62.7) | 65.9 (65.2, 66.5) | 71.2 (70.3, 72.0) | <0.001 |
| 2011-2012 | 67.0 (65.3, 68.6) | 67.7 (65.4, 70.1) | 61.7 (60.3, 63.2) | 65.6 (63.8, 67.3) | 71.6 (69.4, 73.7) | <0.001 |
| 2013-2014 | 66.9 (65.9, 68.0) | 67.3 (66.0, 68.6) | 62.1 (61.2, 63.0) | 66.9 (65.8, 68.0) | 72.0 (70.6, 73.5) | <0.001 |
| 2015-2016 | 67.1 (65.9, 68.4) | 68.1 (66.8, 69.4) | 62.4 (60.7, 64.1) | 64.9 (63.7, 66.0) | 70.8 (69.1, 72.5) | <0.001 |
| 2017-2020 | 67.0 (65.8, 68.1) | 67.7 (66.2, 69.2) | 61.9 (60.5, 63.4) | 66.0 (65.0, 67.0) | 70.6 (69.0, 72.2) | <0.001 |
| **Prevalence of low CVH (<50)** |  |  |  |  |  |  |
| Total | 13.6 (12.7, 14.6) | 12.8 (11.6, 14.1) | 20.7 (19.2, 22.2) | 14.3 (13.0, 15.6) | 7.3 (6.0, 8.5) | <0.001 |
| 2011-2012 | 13.8 (11.6, 16.1) | 13.0 (10.0, 16.7) | 20.5 (16.9, 24.7) | 14.7 (11.5, 18.5) | 8.0 (5.5, 11.6) | 0.007 |
| 2013-2014 | 13.5 (12.3, 14.7) | 12.9 (11.1, 15.0) | 21.7 (19.4, 24.3) | 12.0 (9.5, 15.1) | 7.5 (4.6, 12.1) | <0.001 |
| 2015-2016 | 13.6 (11.2, 15.9) | 12.5 (9.8, 15.8) | 19.3 (16.2, 22.7) | 16.7 (14.1, 19.7) | 6.0 (3.9, 9.1) | <0.001 |
| 2017-2020 | 13.6 (12.0, 15.2) | 12.9 (10.8, 15.3) | 21.1 (18.2, 24.2) | 13.9 (12.1, 16.1) | 7.5 (5.7, 9.8) | <0.001 |

Data were presented as survey weighted mean and 95% CI.

**eTable 3. Each Component of Life’s Essential 8 Score by Race and Ethnicity and Survey Year.**

|  | **Total** | **White** | **Black** | **Hispanic** | **Asian** | ***P* value** |
| --- | --- | --- | --- | --- | --- | --- |
| **Diet** |  |  |  |  |  |  |
| Total | 46.8 (45.6, 47.9) | 47.4 (45.9, 48.9) | 39.2 (37.6, 40.9) | 46.4 (44.9, 47.8) | 57.4 (55.2, 59.5) | <0.001 |
| 2011-2012 | 47.7 (45.5, 49.9) | 49.1 (45.9, 52.3) | 39.2 (34.7, 43.7) | 44.9 (41.7, 48.1) | 55.4 (52.0, 58.8) | <0.001 |
| 2013-2014 | 46.5 (44.9, 48.1) | 46.7 (44.4, 49.1) | 39.9 (37.8, 41.9) | 47.1 (44.2, 49.9) | 55.5 (51.2, 59.9) | <0.001 |
| 2015-2016 | 46.6 (44.6, 48.6) | 47.0 (44.6, 49.5) | 38.8 (34.8, 42.8) | 46.1 (43.7, 48.4) | 57.1 (52.8, 61.4) | <0.001 |
| 2017-2020 | 46.8 (44.5, 49.2) | 47.0 (43.8, 50.2) | 39.1 (36.6, 41.6) | 46.9 (44.0, 49.7) | 59.5 (55.7, 63.4) | <0.001 |
| **Activity** |  |  |  |  |  |  |
| Total | 50.7 (49.1, 52.2) | 53.2 (51.1, 55.3) | 45.5 (43.5, 47.5) | 43.7 (42.2, 45.2) | 52.3 (49.3, 55.2) | <0.001 |
| 2011-2012 | 50.9 (46.8, 55.0) | 54.2 (48.6, 59.7) | 44.8 (40.4, 49.1) | 40.8 (37.2, 44.4) | 51.7 (46.9, 56.5) | 0.003 |
| 2013-2014 | 48.0 (45.3, 50.7) | 49.1 (45.8, 52.3) | 44.5 (40.5, 48.5) | 44.3 (42.3, 46.3) | 53.8 (48.6, 58.9) | 0.006 |
| 2015-2016 | 52.2 (48.5, 55.8) | 55.5 (51.9, 59.2) | 47.7 (42.3, 53.0) | 43.5 (40.3, 46.8) | 47.7 (43.1, 52.4) | <0.001 |
| 2017-2020 | 51.2 (48.7, 53.8) | 53.8 (50.1, 57.4) | 45.3 (42.4, 48.2) | 45.0 (42.6, 47.5) | 54.4 (48.9, 59.9) | <0.001 |
| **Nicotine Exposure** |  |  |  |  |  |  |
| Total | 70.9 (69.8, 72.0) | 69.6 (68.1, 71.1) | 69.0 (67.0, 70.9) | 75.3 (74.1, 76.5) | 84.8 (83.1, 86.5) | <0.001 |
| 2011-2012 | 72.0 (69.9, 74.1) | 70.2 (67.5, 73.0) | 71.9 (67.8, 75.9) | 75.4 (73.0, 77.8) | 84.8 (82.6, 87.0) | <0.001 |
| 2013-2014 | 70.2 (67.8, 72.5) | 68.3 (64.5, 72.2) | 65.8 (63.2, 68.4) | 76.4 (74.0, 78.8) | 83.5 (80.8, 86.1) | <0.001 |
| 2015-2016 | 71.0 (68.7, 73.3) | 69.5 (66.4, 72.6) | 68.1 (65.2, 71.0) | 74.3 (72.4, 76.2) | 84.3 (80.3, 88.3) | <0.001 |
| 2017-2020 | 71.9 (69.8, 74.1) | 70.1 (67.4, 72.8) | 69.7 (65.6, 73.8) | 75.2 (72.9, 77.5) | 85.9 (82.5, 89.3) | <0.001 |
| **Sleep Health** |  |  |  |  |  |  |
| Total | 84.3 (83.7, 85.0) | 86.3 (85.5, 87.1) | 75.9 (74.9, 76.9) | 82.7 (81.9, 83.6) | 86.4 (85.4, 87.5) | <0.001 |
| 2011-2012 | 82.6 (81.1, 84.1) | 83.7 (81.9, 85.6) | 75.0 (72.6, 77.4) | 82.5 (80.6, 84.4) | 85.0 (83.0, 87.0) | <0.001 |
| 2013-2014 | 83.1 (82.0, 84.3) | 84.8 (83.5, 86.0) | 74.3 (72.2, 76.4) | 82.0 (80.1, 83.8) | 86.2 (84.5, 87.9) | <0.001 |
| 2015-2016 | 86.7 (85.7, 87.7) | 89.0 (88.0, 89.9) | 78.0 (76.2, 79.8) | 83.6 (82.3, 85.0) | 87.0 (84.5, 89.5) | <0.001 |
| 2017-2020 | 85.0 (83.8, 86.3) | 87.1 (85.5, 88.7) | 76.2 (74.6, 77.8) | 82.8 (81.3, 84.2) | 87.0 (85.1, 88.9) | <0.001 |
| **Body Mass Index** |  |  |  |  |  |  |
| Total | 58.2 (57.3, 59.2) | 60.2 (58.8, 61.5) | 52.1 (50.9, 53.4) | 54.6 (53.4, 55.8) | 59.4 (57.7, 61.1) | <0.001 |
| 2011-2012 | 61.4 (59.3, 63.6) | 63.6 (60.8, 66.5) | 52.6 (50.4, 54.7) | 57.2 (54.4, 59.9) | 65.0 (61.1, 68.9) | <0.001 |
| 2013-2014 | 58.9 (57.3, 60.5) | 60.4 (58.7, 62.1) | 51.8 (49.1, 54.5) | 56.4 (53.2, 59.5) | 63.5 (60.7, 66.3) | <0.001 |
| 2015-2016 | 58.1 (55.8, 60.4) | 59.9 (57.3, 62.5) | 53.7 (50.6, 56.7) | 52.8 (50.0, 55.5) | 61.1 (59.1, 63.1) | 0.002 |
| 2017-2020 | 56.0 (54.3, 57.8) | 57.9 (55.2, 60.7) | 51.2 (49.3, 53.1) | 53.3 (51.7, 54.9) | 53.4 (51.3, 55.5) | 0.008 |
| **Blood Glucose** |  |  |  |  |  |  |
| Total | 85.2 (84.6, 85.8) | 87.3 (86.5, 88.0) | 79.1 (78.1, 80.0) | 82.7 (81.6, 83.7) | 82.5 (80.9, 84.2) | <0.001 |
| 2011-2012 | 85.9 (84.8, 87.0) | 87.8 (86.1, 89.5) | 78.3 (75.8, 80.8) | 83.6 (81.8, 85.5) | 84.2 (80.9, 87.6) | <0.001 |
| 2013-2014 | 86.4 (85.7, 87.2) | 88.1 (87.2, 89.0) | 80.5 (78.9, 82.1) | 84.8 (83.4, 86.1) | 83.4 (80.4, 86.3) | <0.001 |
| 2015-2016 | 84.3 (82.6, 86.1) | 86.5 (84.8, 88.2) | 78.7 (76.5, 80.8) | 79.9 (76.5, 83.3) | 83.8 (80.8, 86.8) | <0.001 |
| 2017-2020 | 84.8 (83.8, 85.8) | 86.9 (85.4, 88.5) | 78.9 (77.2, 80.5) | 82.6 (81.3, 83.9) | 80.4 (77.6, 83.2) | <0.001 |
| **Blood Lipids** |  |  |  |  |  |  |
| Total | 67.4 (66.5, 68.3) | 66.5 (65.4, 67.7) | 73.6 (72.5, 74.7) | 66.6 (65.3, 67.9) | 67.5 (66.0, 69.0) | <0.001 |
| 2011-2012 | 63.7 (61.8, 65.6) | 62.5 (60.3, 64.8) | 69.1 (67.2, 71.0) | 63.6 (60.9, 66.4) | 67.9 (65.1, 70.7) | <0.001 |
| 2013-2014 | 67.5 (65.9, 69.1) | 66.9 (64.9, 68.8) | 73.7 (71.3, 76.2) | 65.5 (62.9, 68.0) | 67.9 (65.0, 70.7) | 0.003 |
| 2015-2016 | 67.6 (65.9, 69.2) | 66.7 (64.8, 68.6) | 74.1 (71.9, 76.2) | 66.4 (64.7, 68.0) | 67.8 (65.1, 70.6) | <0.001 |
| 2017-2020 | 69.5 (67.9, 71.1) | 68.8 (66.5, 71.0) | 75.9 (73.8, 78.0) | 68.9 (66.5, 71.4) | 66.9 (64.0, 69.8) | <0.001 |
| **Blood Pressure** |  |  |  |  |  |  |
| Total | 70.7 (70.0, 71.5) | 70.8 (69.7, 71.9) | 63.2 (62.1, 64.3) | 74.7 (73.6, 75.8) | 73.6 (72.3, 74.9) | <0.001 |
| 2011-2012 | 71.0 (69.0, 73.0) | 70.5 (68.0, 73.1) | 64.3 (61.3, 67.4) | 76.6 (73.4, 79.7) | 76.2 (73.5, 78.9) | <0.001 |
| 2013-2014 | 71.4 (70.1, 72.7) | 71.4 (69.4, 73.3) | 63.1 (61.0, 65.2) | 76.7 (74.8, 78.7) | 74.8 (71.8, 77.8) | <0.001 |
| 2015-2016 | 69.4 (68.0, 70.7) | 69.2 (67.6, 70.9) | 63.9 (61.6, 66.1) | 72.1 (70.0, 74.3) | 73.8 (70.6, 77.0) | <0.001 |
| 2017-2020 | 71.0 (69.5, 72.4) | 71.8 (69.5, 74.1) | 62.1 (60.2, 64.0) | 74.1 (72.2, 75.9) | 71.3 (69.5, 73.1) | <0.001 |

**eTable 4. Z-Scores for Racial Differences in Life's Essential 8 Components.**

|  | **Diet** | **Physical Activity** | **Nicotine Exposure** | **Sleep Health** | **Body Mass Index** | **Blood Glucose** | **Blood Lipids** | **Blood Pressure** |
| --- | --- | --- | --- | --- | --- | --- | --- | --- |
| **2011-2012** |  |  |  |  |  |  |  |  |
| Black-White | -1.18 | -1.11 | 0.43 | -1.02 | -1.35 | -1.13 | 1.13 | -0.66 |
| Hispanic-White | -0.38 | -1.67 | 0.93 | 0.03 | -0.70 | -0.38 | 0.36 | 1.06 |
| Asian-White | 1.09 | -0.14 | 2.25 | 0.39 | 0.40 | -0.30 | 0.96 | 1.00 |
| **2017-2020** |  |  |  |  |  |  |  |  |
| Black-White | -0.88 | -0.97 | 0.21 | -1.33 | -0.72 | -0.91 | 1.32 | -1.15 |
| Hispanic-White | 0.26 | -1.01 | 1.01 | -0.37 | -0.41 | -0.37 | 0.29 | 0.60 |
| Asian-White | 2.12 | 0.37 | 2.59 | 0.25 | -0.39 | -0.68 | -0.01 | 0.20 |

The Z-scores are calculated by standardizing values of all components across all racial and ethnicity groups.
